# Comparing Cigarette-Cue Attentional Bias Between People With HIV/AIDS and People With Opioid Use Disorder Who Smoke

**DOI:** 10.1101/2022.09.30.22280501

**Authors:** Gopalkumar Rakesh, Joseph L Alcorn, Rebika Khanal, Seth S Himelhoch, Craig R Rush

**Affiliations:** Department of Psychiatry, College of Medicine, University of Kentucky, Lexington, KY,USA; Department of Psychology, College of Arts & Sciences, University of Kentucky, Lexington, KY,USA; Department of Behavioral Sciences, College of Medicine, University of Kentucky, Lexington, KY,USA

**Keywords:** HIV/AIDS, opioid use disorder, cigarettes, attentional bias

## Abstract

**Background:** Special populations like people living with HIV/AIDS (PLWHA) and people with opioid use disorder (OUD) smoke tobacco cigarettes at rates three to four times greater than the general population. Patients with tobacco use disorder exhibit attentional bias (AB) for cigarette cues, which is theorized to be caused by classical conditioning and motivational salience. Eye tracking can quantify this bias by measuring fixation time (FT) on cigarette and matched neutral cues, which consequently helps calculate an AB score. Although previous studies have measured this bias in people who smoke without any other comorbid conditions, no study has measured this bias in special populations.

**Methods:** We analyzed eye tracking data collected from participants in two randomized clinical trials (RCTs). The study protocols that recruited participants were approved by Medical Institutional Review Board (IRB) at University of Kentucky and were registered on clinical trials.gov (NCT05049460, NCT05295953). We compared FT and AB score towards cigarette cues between PLWHA and people with OUD who smoke, using a visual probe task and Tobii Pro Fusion eye tracker. We used two cigarette cue types, one encompassing people smoking cigarettes and the other consisting of cigarette paraphernalia. We used two cue presentation times, 1000 and 2000 milliseconds (ms).

**Results:** Cues of people smoking cigarettes elicited greater AB than cues of cigarette paraphernalia across both subject groups when cues were presented for 2000 ms, but not 1000 ms. PLWHA who smoke exhibited greater AB for cues of people smoking cigarettes than cigarette paraphernalia when presented for 2000 ms compared to people with OUD who smoke.

**Conclusion:** The addition of social cues potentiates cigarette cue AB, based on cue type and stimulus presentation time. Understanding the neurobiology of this relationship can help design treatments that can target AB and prevent relapse.

## 1. Introduction

Smoking is the leading preventable cause of death and disease in the United States^1^. Fourteen percent (14%) of adults in the United States smoke ^2^. People living with HIV/AIDS (PLWHA) smoke at nearly three to four times the rate of the general population ^3^. Smoking in PLWHA is also associated with increased incidence of comorbid substance use disorders, low adherence to antiretroviral therapy (ART) medications and consequently increased mortality and poor quality of life ^4,5^.

One additional group of individuals with disproportionally high rates of cigarette use are individuals with opioid use disorder (OUD). Approximately 74 – 94% of patients with OUD smoke ^6^. The mechanisms for high smoking rates in individuals with OUD are unknown. Two proposed pharmacological mechanisms are crosstolerance between nicotine and opioids ^7,8^ and nicotine ameliorating pain via alpha-4-beta-2 acetylcholine receptors agonism (α4β2) in individuals with OUD ^8^. In addition, medications used to treat OUD (e.g., methadone and suboxone) are associated with increased smoking rates ^9-12^.

Patients with a current or previous history of tobacco use disorder demonstrate an attentional bias (AB) for cigarette cues when exposed to cigarette and neutral cues ^13-15^, whereas those without a history of tobacco use disorder do not. AB to cigarette cues has also been shown to be associated with increased craving and propensity for relapse in people who smoke cigarettes ^16^. Eye-tracking measures can be utilized to quantify the differential allocation of attention to salient cues (e.g., cigarettes) as compared neutral cues ^17,18^. Eye-tracking measures such as the visual probe task utilize fixation time (FT), measured in milliseconds towards salient cues and neutral cues as an index of AB ^19,20^. AB have been shown to be consistent over longitudinal periods of time when measured in subjects with substance use disorders ^20^. Plausible modulators of AB include abstinence, the presence of withdrawal, craving for the substance of choice, sensory modality, duration of salient cue presentation, and emotional valence of cues ^17,21^.

Previous work quantified cigarette-cue AB via eye-tracking measures with cue presentation times ranging from 200 to 2000 ms ^22-27^. All these studies compared people with and without tobacco use disorder. No previous studies have examined AB to cigarette cues in populations with higher rates of smoking such as PLWHA or people with OUD.

The present study sought to compare cigarette-cue AB between PLWHA who smoke tobacco cigarettes and people with OUD who smoke tobacco cigarettes. In addition, previous studies have not compared how cigarette-cue AB differs based on cue type (people smoking cigarettes versus cigarette paraphernalia) but have suggested it as a potential area to explore ^21^. We used two cue types-people smoking cigarette cues and cigarette paraphernalia in separate trials. For each cue type, we used cue presentation times of 1000 milliseconds (ms) and 2000 ms. Previous studies demonstrated greater AB at longer cue durations ^22,23^. The primary hypothesis of the present study was cigarette-cue AB towards both cue types would be significantly greater with presentation times of 2000 ms compared to 1000 ms. Given higher substance use comorbidity and disease burden in PLWHA who smoke, we also hypothesized PLWHA who smoke would show greater cigarette-cue AB than people with OUD who smoke ^5^.

## 2. Methods

### 2.1 Participants

We analyzed eye tracking data collected from participants in two randomized clinical trials (RCTs). These participants were recruited in two RCTs comparing effects of active versus sham transcranial magnetic stimulation (TMS) on cigarette-cue attentional bias. We analyzed data from ten PLWHA who smoke tobacco cigarettes (1 female, 9 male) and ten participants with OUD who smoke tobacco cigarettes (6 females, 4 males). Both sets of participants came from two clinics affiliated with the University of Kentucky Medical Center, the data we analyzed was acquired at baseline. Participants were randomized to receive either active or sham TMS.

### 2.2. Procedures

The study protocols that recruited participants were approved by Medical Institutional Review Board (IRB) at University of Kentucky and were registered on clinical trials.gov (NCT05049460, NCT05295953). Prior to study day, all participants completed a phone screen to determine eligibility. Once deemed eligible for the study, participants were instructed to abstain from smoking cigarettes or using other substances for two hours prior to start of experimental session.

On study day, all participants completed an IRB approved informed consent. Demographic information, medical history, and current and past use of tobacco were then assessed. Smoking abstinence was corroborated with carbon monoxide (CO) measurement using a Smokerlyzer Breath CO Monitor (Bedfont Scientific Ltd., Rochester, England). Blood alcohol level (BAL) was also recorded using a Breathalyzer (Alco-Sensor FST, St. Louis, MO, USA). Participants were required to have a CO level less than 10 ppm and a BAL of 0.000% to proceed with study procedures. Participants completed Fagerstrom Test for Nicotine Dependence (FTND) and four sets of AB trials. Two sets of AB trials measured AB for cues encompassing people smoking cigarette versus neutral cues and two sets of AB trials measured AB for cigarette paraphernalia cues versus neutral cues.

### 2.3. Visual Probe Task

Attentional Bias was measured using the visual probe task based on ^28^ study. Fixation data were collected using Tobii Pro Fusion 120 Hz eye tracker (Tobii Technology, Sweden). For each AB trial, images of a cigarette (C) and a matched neutral (N) cue were presented on a laptop screen, 3 cm apart. Upon offset of the image pairs, a visual probe (X) appeared on either the left side or right side of the screen, in the same location as one of the previously presented images. Firstly, twenty trials showed images of cues encompassing people smoking cigarettes and matched neutral cues (C-N) for 1000 ms. Subsequently, additional twenty trials showed the same image pairs for 2000 ms. Secondly, twenty trials showed images of cues encompassing cigarette paraphernalia and matched neutral cues (C-N) for 1000 ms. Subsequently, additional twenty trials showed the same image pairs for 2000 ms.

For each set of twenty cigarette-cue AB trials, twenty filler trials consisting of twenty pairs of additional neutral cue images (N-N) were intermixed to generate a total of 40 trials. Filler trials were also presented for 1000 or 2000 ms, depending on presentation times of C-N cue pairs. Intervals between C-N and N-N cue pairs were 1000 or 2000 ms, contingent on cue presentation time. Please see Supplementary material figure 1 for a visual of cue presentation.

Cigarette cue images were taken from a validated database (Smoking Cue Database [SmoCuDa]) ^29^. The database divided 250 image cues related to smoking, into eighteen categories based on similarities in cue description. Each cue image in SmoCuDa was rated by 40 participants who smoked, on ratings of ‘urge to smoke’, ‘valence’ and ‘arousal’ between 1 and 100. For cues of people smoking cigarette cues, we chose the first twenty relevant images in the database, ranked by urge to smoke from highest to lowest. Cues were counterbalanced for sex, ethnicity, and affective content. For cues encompassing cigarette paraphernalia, we chose the first twenty relevant images ranked by urge to smoke from highest to lowest.

### 2.4. Data analytic strategy

A trial level bias score (TLBS) was calculated for every trial by FT on cigarette cue minus FT on neutral cue. We calculated averages of trial level FT for cigarette cues, trial level FT for neutral cues, and trial level AB score for cigarette cues for each set of cues separately for 1000 ms and 2000 ms of cue presentation times in both participant groups. Any trial wherein the FT on both cigarette and neutral cue were both null was deemed an outlier and removed. As part of cleaning the data, we removed 101 outlier observations which had null values for FT on cigarette and neutral cues. This reduced total number of total trials from 1600 to 1459.

All trial level FT for cigarette cues and respective AB scores were entered into separate linear fixed effects regression models. Outcome variables in one model was FT on cigarette cues and AB score in the other model. Predictor variables in both models included age, gender, subject group (PLWHA who smoke versus OUD who smoke), cigarette cue type (people smoking cigarette cues and neutral cues versus cigarette paraphernalia cues and neutral cues), cue presentation time (1000 ms versus 2000 ms of presentation time for cue pairs) and FTND scores. All these predictor variables were used as fixed effects and subject ID was the grouping variable. We performed all analyses using Matlab version R2022a (Natick, Massachussetts: The Mathworks Inc.)

## 3. Results

An independent-samples t-test revealed statistically significant difference in age between the two groups. (t(18) =3.39, p=0.003). The mean age in the PLWHA who smoke group was 44.6 years (SEM = ±3.14) and 33 years (SEM = ±1.76) for people with OUD who smoke. A separate independent samples t-test revealed no statistically significant difference in FTND scores (t(18) = 0.91, p = 0.37). The respective mean FTND scores for the PLWHA and OUD groups were 6.10 and 7 (SEMs = ± 0.87 and 0.56, respectively). A chi-square test revealed that a significant difference in the sex distribution (χ2(1) = 17.35, p = 0.02). Given our limited sample, we did not include age or gender as predictor variables in our regression models. FT was highest in the PLWHA group presented with cues of people smoking cigarettes and neutral cues, with stimulus presentation time of 2000 ms (Figure 1 and Table 1).

**Figure 1.**
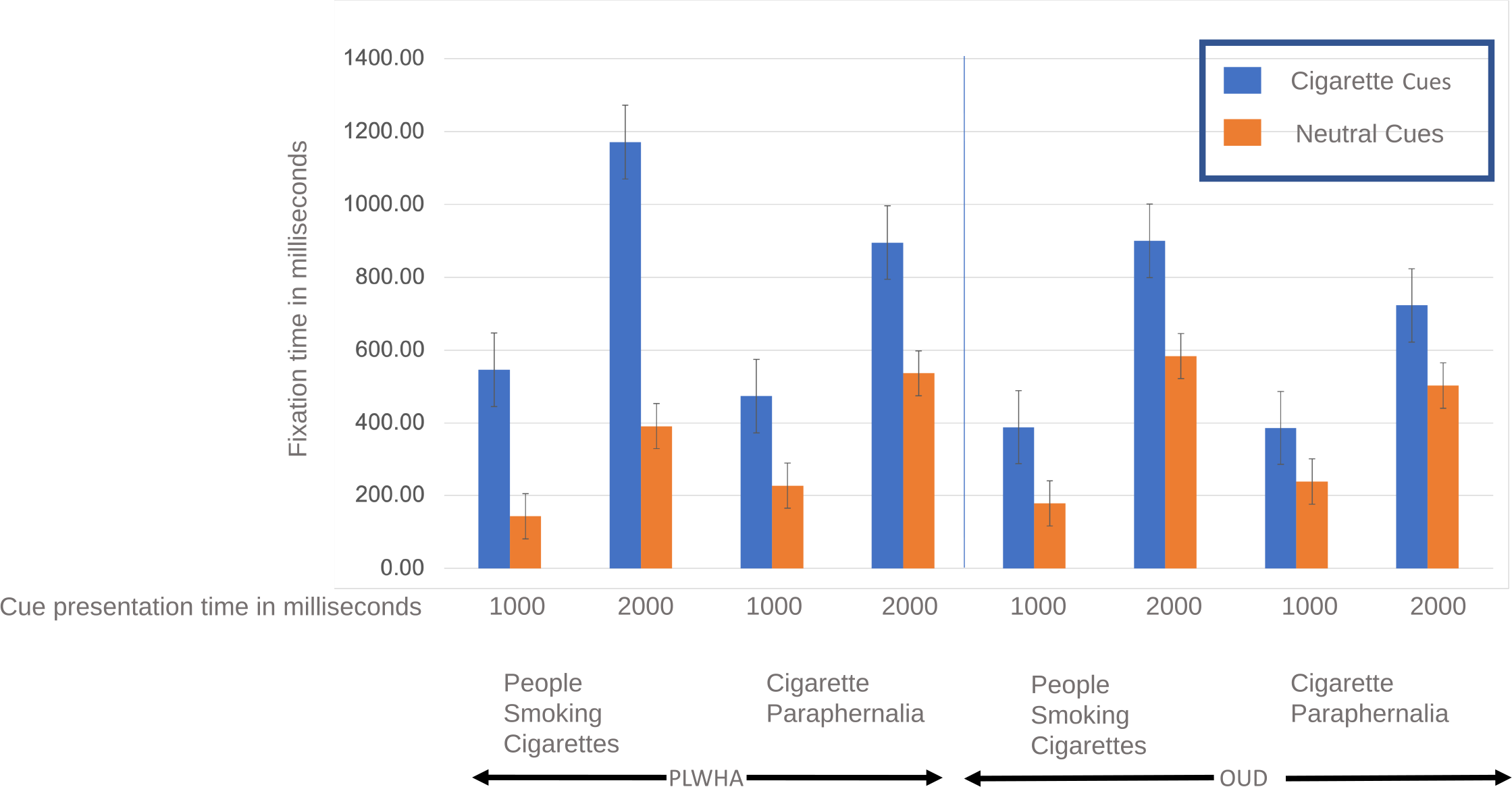
Fixation Time on Cigarette and Neutral Cues. showcases bars encapsulating fixation time (FT) in milliseconds for cigarette cues and neutral cues. The X-axis lists cue presentation times (1000 milliseconds versus 2000 milliseconds), cue type (people smoking cigarettes with matched neutral cues versus cigarette paraphernalia with matched neutral cues) and subject group (PLWHA who smoke versus patients with OUD who smoke). The Y-axis shows fixation time in milliseconds. Bars in blue represent FT on cigarette cues and bars in orange represent FT on neutral cues. PLWHA who smoke had higher FT on both cigarette cue types when presented for 2000 milliseconds, compared to people with OUD who smoke. Across both subject groups, cues of people smoking cigarettes elicited significantly higher FT than cues of cigarette paraphernalia Fixation Time on Cigarette and Neutralwhen presented for 2000 milliseconds

**Table 1.**
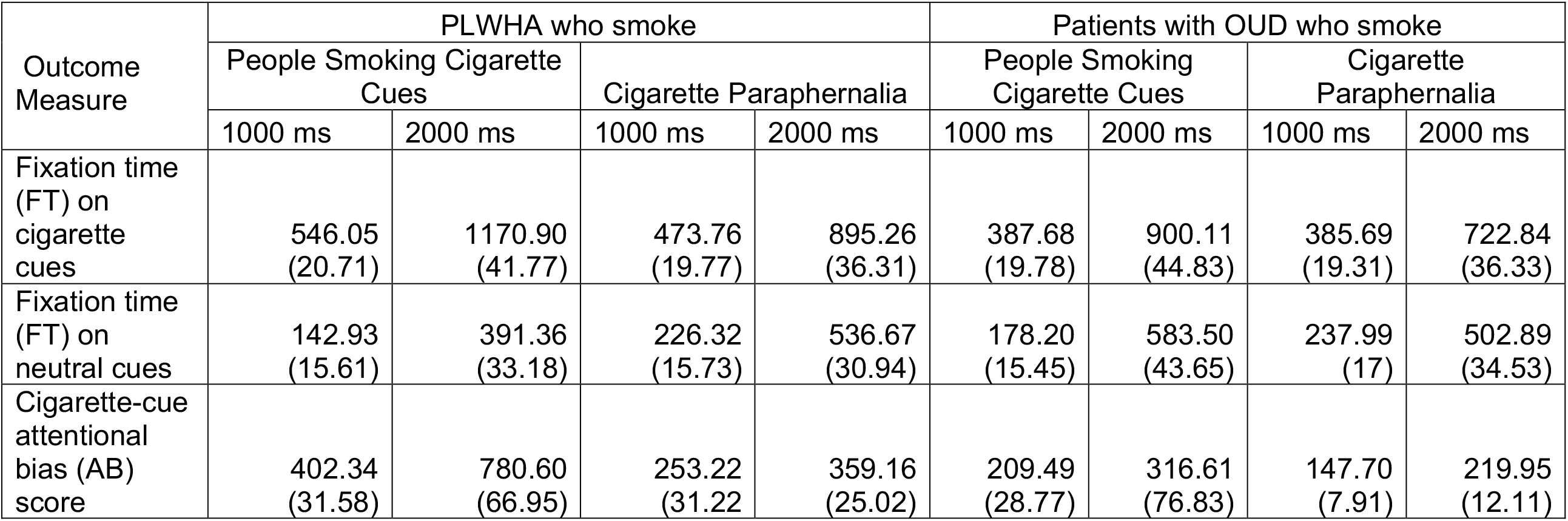
Fixation Time (FT) and Cigarette-cue AB score Mean values in cells and standard error of mean in parentheses

Table 2 shows a linear fixed effects regression model with FT on cigarette cues as outcome. Cues of people smoking cigarettes elicited greater FTs than cues of cigarette paraphernalia across both subject groups (Table 2). This is corroborated by significant association between FT and cue type (t(1446) = 1.96, p = 0.05) and no interaction between subject group and cue type (t(1446)=-0.09, p = 0.93)(Table 2). Across both subject groups, cues of people smoking cigarettes showed significantly higher FT than cigarette paraphernalia cues with 2000 ms but not with 1000 ms, as reflected by the significant association between cue presentation time and FT (t(1446) = 11.43, p < 0.00001) and significant interaction between cue type and cue presentation time (t(1446) = -3.28, p = 0.001) (Table 2). Although PLWHA showed greater FT for cigarette cues than patients with OUD (Table 1) this was not significant when controlling for other predictors (Table 2).

**Table 2.**
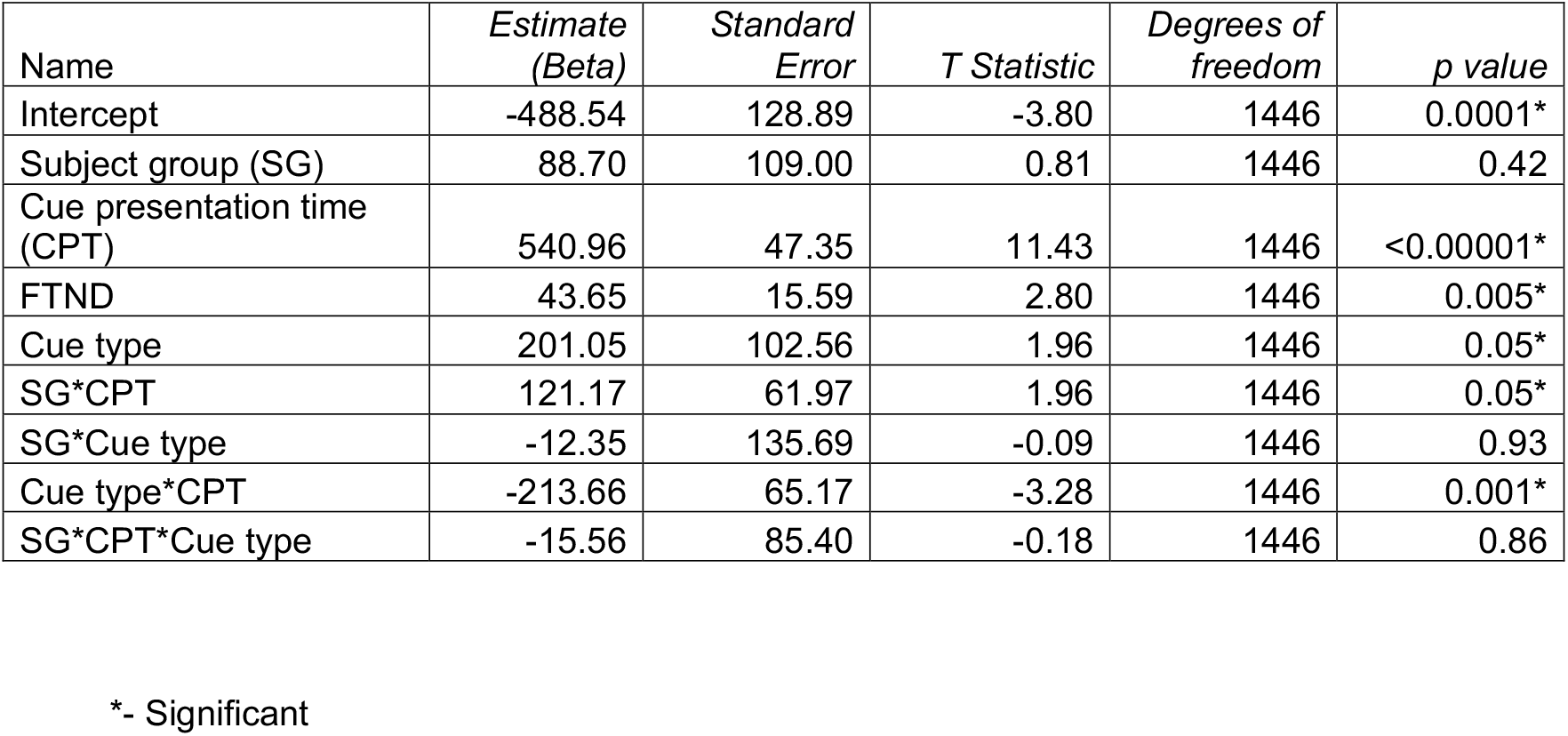
Linear Fixed Effects Model Fixation Time (FT) on Cigarette Cues as Outcome Measure Fixation on cigarette cues (milliseconds) ∼ 1 + subject group (SG)(PLWHA who smoke versus OUD who smoke) + cue presentation time (CPT) (1000 milliseconds versus 2000 milliseconds of cue presentation) + Fagerstrom test for nicotine dependence (FTND) scores + cue type (cues of people smoking cigarette versus cigarette paraphernalia cues) + SG * cue type + SG * CPT + cue type * CPT + SG * CPT * Cue type + 1|Subject ID. (R^2^ = 0.44)

Table 3 shows a linear fixed effects regression models with cigarette-cue AB score as the outcome measure. Neither cue type nor cue presentation time showed significant associations with cigarette-cue AB score (Table 3). PLWHA who smoke exhibited significantly greater cigarette-cue AB score for people smoking cigarette cues when presented for 2000 ms instead of 1000 ms compared to people with OUD who smoke, as shown by a significant three-way interaction between diagnostic group, cue presentation time and cue type (t(1446) = -1.98, p = 0.05) (Table 3).

**Table 3.** Linear fixed effects model Cigarette-cue AB score as Outcome Measure Cigarette-cue AB score ∼ 1 + subject group (SG)(PLWHA who smoke versus OUD who smoke) + cue presentation time (CPT) (1000 milliseconds versus 2000 milliseconds of cue presentation) + Fagerstrom test for nicotine dependence (FTND) scores + cue type (cues of people smoking cigarette versus cigarette paraphernalia cues) + SG * cue type + SG * CPT + cue type * CPT + SG * CPT * Cue type + 1|Subject ID. (R^2^ = 0.18)

| Name | *Estimate*<br>*(Beta)* | *Standard*<br>*Error* | *T Statistic* | *Degrees of*<br>*freedom* | *p value* |
| --- | --- | --- | --- | --- | --- |
| Intercept | -73.75 | 185.95 | -0.40 | 1446 | 0.69 |
| Subject group (SG) | -159.11 | 177.07 | -0.90 | 1446 | 0.37 |
| Cue presentation time (CPT) | 101.89 | 79.40 | 1.28 | 1446 | 0.2 |
| FTND | 33.29 | 20.18 | 1.65 | 1446 | 0.1 |
| Cue type | -111.16 | 171.84 | -0.65 | 1446 | 0.52 |
| SG*CPT | 311.74 | 104.99 | 2.97 | 1446 | 0.003* |
| SG*Cue type | 277.83 | 229.86 | 1.21 | 1446 | 0.23 |
| Cue type*CPT | -3.87 | 109.40 | -0.04 | 1446 | 0.97 |
| SG*CPT*Cue type | -286.91 | 144.80 | -1.98 | 1446 | 0.05* |

\*-Significant

## 4. Discussion

To the best of our knowledge, this is the first study to compare AB for cigarette cues between these populations who smoke. In alignment with our results, the SmoCuDa database showed greater urge to smoke and valence for cues of people smoking cigarettes than cigarette paraphernalia ^29^. Two other studies did not show any association between affective nature of cues and FT for cigarette cues in people who smoke ^22,26^.

Comparing patients with tobacco use disorder to controls, four notable studies showed an AB towards cigarette cues using response times and three studies showed an AB towards cigarette cues using FT via eye tracking ^22-27,30^. These seven studies used variable cue presentation times: 500 ms ^27,30^, 1000 ms^25^, only 2000 ms ^24^, 200 and 2000 ms ^22^, 500 ms and 2000 ms ^23^. Except for one study which showed an AB when cues were presented for 500 ms ^30^, all other studies showed an AB towards cigarette cues were presented for 1000 and 2000 ms, but not for 200 or 500 ms ^22-27^. Although previous studies have compared cue presentation times of 500 ms and 2000 ms, ours is the first to compare 1000 and 2000 ms.

Although FT on cigarette cues could have some conceptual overlap with cue reactivity, the goal with FT measurement is not to elicit craving in response to cue presentation ^17,18^. The neurobiological basis of cigarette-cue reactivity suggests dopaminergic sensitization in the mesolimbic pathway, decrease in central executive network (CEN) functional connectivity, and increase in salience network functional connectivity^14,21^. The neurobiological mechanism for cue modulation on AB is unknown ^31^. Future studies examining the neural correlates of cigarette-cue AB are warranted.

## 5. Conclusions and limitations

The sample size was small limiting generalizability. We found greater cigarette-cue AB score when cues were presented for 2000 ms compared to 1000 ms for both special populations who smoke at elevated rates. Cues comparing people smoking cigarettes and equivalent neutral cues elicited greater FT in both populations than cues of cigarette paraphernalia with corresponding neutral cues. PLWHA who smoke showed greater cigarette-cue AB score than people with OUD who smoke for cues of people smoking cigarettes when presented for 2000 ms. These preliminary results do support exploring neural correlates of these cue presentations, to identify a valid behavioral marker of cigarette use and relapse.

## Data Availability

Our study data is available on Mendeley. The DOI is http://dx.doi.org/10.17632/g6yzzr34sx.1

http://dx.doi.org/10.17632/g6yzzr34sx.1

## Acknowledgements

This work was supported by the National Institutes of Health (grant numbers CA225419, DA035200) and University of Kentucky College of Medicine.

## Data availability statement

Our study data is available on Mendeley. The DOI is http://dx.doi.org/10.17632/g6yzzr34sx.1

---

**Supplementary Figure 1.**
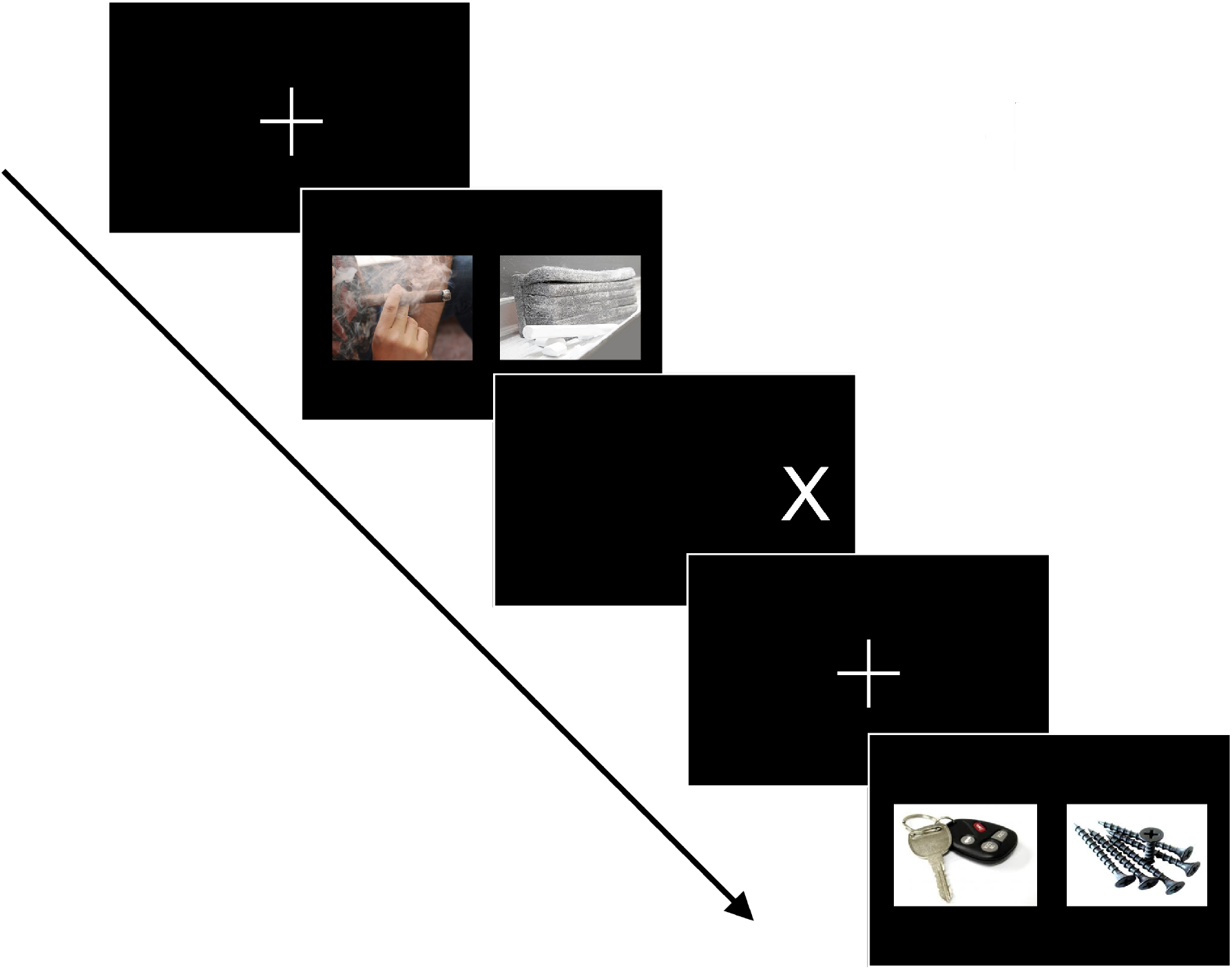
Cue presentation to measure fixation time and calculate cigarette-cue AB score

